# Is linear regression sufficient for normative psychological assessment? A comparison between OLS, GAM, and GAMLSS across seven public reference datasets

**DOI:** 10.64898/2026.09.16.26363213

**Authors:** Marta Arbizu Gómez, Pablo Rodriguez-Royo, Elina Maltseva, Carolina Sastre-Barrios, Jorge M. Corada, César Ortea Suárez, Íñigo Fernández de Pierola, Marcos Ríos-Lago, Jesús M. Cortés

## Abstract

**Background:** Normative Z-scores are essential for interpreting individual cognitive performance relative to healthy reference populations. Traditional norms often compare individuals within predefined groups, such as age bands. This is useful and easy to interpret, but it can lose information because variables such as age change continuously and because age, sex, and education may contribute jointly to performance. Regression-based norms estimate the expected score for each individual from their specific covariates and then express the difference between the observed and expected score as a normative Z-score. Therefore, the key methodological question is not simply whether a more complex model can be fitted, but whether that added complexity changes the Z-score in a meaningful way.

**Methods:** We implemented a comparative framework and fitted ordinary least squares (OLS), generalized additive models (GAM), and generalized additive models for location, scale, and shape (GAMLSS) to the seven public reference datasets distributed in the NormData R package. The datasets span processing speed, verbal memory, verbal fluency, academic achievement, personality, and anxiety. For each dataset we compared each regression model with the classic subgroup norm, and then compared the regression models with one another. Metrics included Pearson r, residual RMSE, and the percentage of participants whose model-based Z-score agreed with the classic Z-score.

**Results:** All three models reproduced the classic Z-scores closely (r = 0.964-1.000). OLS and GAM produced nearly identical Z-scores throughout (r = 0.996-1.000), indicating that the observed age trajectories were smooth enough for a quadratic term to capture. GAMLSS diverged from OLS mainly in datasets with non-constant residual variance (Fluency, sigma-ratio = 1.79; TMAS, sigma-ratio = 1.50), where modeling the scale parameter recovered dispersion differences that the classic subgroup method encodes implicitly.

**Conclusions:** Across seven heterogeneous reference datasets, OLS provides a sufficient normative Z-score computation when the sample size is large enough and residual variance is approximately constant. GAM is useful when the mean trajectory is strongly nonlinear; GAMLSS is useful when residual variance changes across covariate profiles. The practical message is deliberately simple: add model complexity only when the data show what that complexity contributes to the individual Z-score.

## 1. Introduction

Neuropsychological and psychological assessment involves the integration of clinical history, behavioural and semiological observations, and standardized test performance, with normative reference data providing an essential framework for interpreting an individual’s cognitive results. Traditionally, normative data have been obtained by partitioning reference samples into demographic subgroups, typically defined by age, sex, and educational level, and then computing group-specific means and standard deviations (Mitrushina et al., 2005; Strauss et al., 2006; Sherman et al., 2022; Tombaugh 2004). In simple terminology, a norm tells us what score is expected for a person with a similar profile. The limitation is that the traditional method needs enough people in every subgroup, and it treats age cut-offs as if they were natural boundaries rather than convenient bins (Crawford & Howell, 1998; Heaton et al., 2004).

Regression-based normative modeling has been proposed as a principled alternative (Zachary & Gorsuch, 1985; Van der Elst et al., 2005, 2006). Expected performance is modeled as a continuous function of demographic covariates, and individual deviations are quantified through standardized residuals. This approach accommodates nonlinear covariate effects, adjusts simultaneously for multiple predictors, and can reduce the sample size required for precise norms (Innocenti et al., 2023). It is the methodological basis of the NormData R package and of a growing applied literature on regression-based norms (Van der Elst, 2024).

Recent work has shown that regression-based norms are useful for modeling raw scores directly rather than transform the observed response for normality (Oltra-Cucarella et al., 2025). Their Spanish older-adult normative studies also illustrate how regression-based norms can be developed for specific populations rather than borrowed from a less appropriate reference group (Calderón-Rubio et al., 2021; Iñesta et al., 2021, 2022). The same need for population-specific norms has been also shown for Latin American, Spanish-speaking, and pediatric samples, where regression and related modern psychometric models are used to adapt interpretation to age, education, sex, language, and cultural context (delCacho-Tena et al., 2023; Fuentes Mendoza et al., 2025; Rivera et al., 2019).

Within the regression-based family, three models of increasing flexibility are available. Ordinary least squares (OLS) regression models the conditional mean as a function of covariates and conventionally assumes constant residual variance. Generalized additive models (GAMs) replace fixed polynomial terms with smooth penalized splines, allowing the data to determine the shape of each covariate effect (Hastie & Tibshirani, 1986; Wood, 2017). Generalized additive models for location, scale, and shape (GAMLSS) extends this further by modeling not only the conditional mean but also the conditional standard deviation as functions of covariates, directly addressing heteroscedasticity (Rigby & Stasinopoulos, 2005).

The key practical question is not whether GAM or GAMLSS can model more complexity than OLS; by construction they can. The question is whether that added flexibility changes the normative Z-score in a way that matters. Here, we make use of a public-dataset for model comparison, and then decide whether model complexity adds information or simply makes the model harder to explain.

## Disclosures

### Preregistration

No part of this study was preregistered. The work is a secondary, methodological analysis of previously collected, publicly available reference datasets; the comparisons between normative models were exploratory and were not based on preregistered confirmatory hypotheses.

### Data, materials, and online resources

All datasets analysed in this study are publicly available in the NormData R package (version 1.1; Van der Elst, 2024, https://doi.org/10.32614/CRAN.package.NormData). No new data were collected. The analysis code that supports the findings (data preprocessing, model fitting, bias correction, and all reported comparisons) is available from the corresponding author upon reasonable request.

### Reporting

This study involved an analysis of existing data rather than new data collection. We report all datasets analysed, all models fitted, all data exclusions, and all metrics computed in the study.

### Ethical approval

Ethical approval was not required for this study. The analyses were conducted exclusively on previously collected, de-identified, publicly available datasets (and, in some cases, simulated data) distributed with the NormData R package; no new data were collected from human participants by the authors, and no identifiable information was accessed.

## 2. Methods

### 2.1 General definition of normative Z-score

For an outcome Y and a vector of demographic covariates X, the normative Z-score for individual i was defined as Eq. (1), where 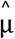 is the estimated expected score and 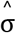 is the estimated residual standard deviation for that profile. Put simply, the score asks how far the person is from what the model expects for someone similar.

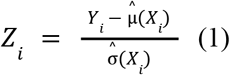

### 2.2 Classic method

The reference sample was partitioned into subgroups defined by the cross-classification of the available demographic factors; continuous age, when present, was binned into 10-year intervals. Within each cell, the mean and standard deviation of Y were computed, and Eq. (2) was applied. A Z-score was computed only when the cell contained at least five observations and a non-zero standard deviation.

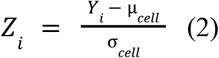

### 2.3 OLS regression

OLS estimated the conditional mean with Eq. (3). Age was mean-centered before forming the quadratic term, and the lowest educational level was used as the reference category. Only covariates significant in univariate screening (p < 0.05) were retained; the quadratic age term was kept only when significant in its own right. The residual standard deviation was a single constant, so OLS standardized residuals according to Eq. (4).

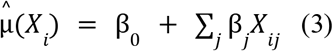

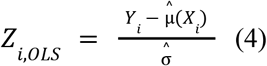

### 2.4 Generalized additive models (GAM)

GAMs modeled the conditional mean with Eq. (5), where *f* _λ_ (age) is a penalized spline estimated from the data and categorical covariates enter as factor terms. The penalty parameter λ controls the trade-off between fit and smoothness, whereby a large λ is useful when the goal is a stable and smooth age trend (avoiding overfitting), whereas a small λ is useful when the data support a more flexible age function that can capture specific nonlinear changes. Here, λ was selected automatically by grid search. The effective degrees of freedom (EDF) of the age spline quantify its complexity: EDF close to 1 indicates a quasi-linear effect, whereas higher EDF indicates more curvature. GAMs were fitted with pyGAM (Servén & Brummitt, 2018).

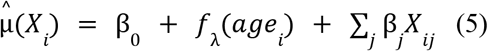

### 2.5 Generalized additive models for location, scale, and shape (GAMLSS)

GAMLSS additionally models the conditional standard deviation as a function of covariates (Rigby & Stasinopoulos, 2005). A Gaussian family was assumed, making the specification directly comparable to OLS and GAM. In the present implementation, the scale model was approximated by a two-stage pyGAM procedure: a GAM for the mean, followed by a GAM fitted to the log of squared residuals to estimate conditional log-variance, iterated with reweighting.

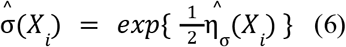

Because the second stage regresses log squared residuals on covariates, a known bias arises under Gaussian errors (Harvey, 1976). The bias term is shown in Equation (7). We therefore also computed a bias-corrected GAMLSS by adding 1.2704 to the log-variance predictor before exponentiation.

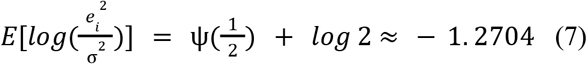

### 2.6 Comparisons and metrics

Two comparisons were conducted for each dataset. First, each model’s Z-scores were correlated with the classic Z-scores (Pearson r over participants with a valid value in both), assessing equivalence with established norms. Second, the regression models were correlated with one another (OLS vs. GAM, OLS vs. GAMLSS, GAM vs. GAMLSS), assessing what the added flexibility contributes. In addition to Pearson r we report residual RMSE on the original test scale and the percentage agreement within 0.25 Z-score units, defined in Equation (8).

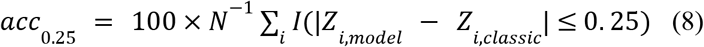

### 2.7 Nonparametric Z-score

The parametric Z-score assumes that model residuals are approximately normally distributed. To avoid making that assumption, we computed a nonparametric rank-based Z-score as the final score for every dataset. Residuals were ranked, converted to empirical percentiles as in Equation (9), and mapped onto the standard normal scale through Equation (10). This preserves the ordering of the residuals while giving valid percentiles even when residuals are skewed.

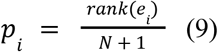

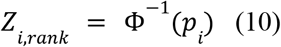

### 2.8 Reference datasets and implementation

The seven datasets distributed with the NormData R package differ in the construct measured and in the recorded covariates. Two contain age, sex, and educational level; two contain only educational level; and three contain only sex. The model and the subgroup scheme were adapted to the covariates available in each dataset. All analyses were implemented in Python using NumPy, pandas, SciPy, and pyGAM in a fully scripted, reproducible pipeline.

## 3. Results

### 3.1 Covariate selection and model complexity

Univariate screening produced a data-driven OLS model for each dataset, with the number of retained predictors ranging from zero (STAS, intercept-only) to five (VLT). For LDST, age and educational level were retained while sex and the quadratic age term were not, yielding a model consistent with the NormData reference materials (Van der Elst, 2024). Rather than keep a procedural p-value table in the main manuscript, we summarize the model complexity and the model-to-model agreement in Table 2, which is more directly connected to the methodological question.

**Table 1.** Public reference datasets used for model comparison. For each dataset, the table reports a brief description of the test and the construct it measures, the outcome (normed variable), the sample size, and the available demographic covariates. All datasets are distributed in the NormData R package (Van der Elst, 2024).

| Dataset | Description (what it is and what it measures) | Outcome | N | Available covariates |
| --- | --- | --- | --- | --- |
| Substitution (LDST) | Letter-Digit Substitution Test, a speeded paper-and-pencil substitution task; measures information-processing / psychomotor speed. | Letter Digit Substitution score | 1,650 | age, sex, education |
| VLT | Verbal Learning Test (Rey's Verbal Learning Test paradigm), word-list learning; measures verbal learning and episodic memory. | Verbal Learning - total recall | 1,460 | age, sex, education |
| Fluency | Semantic (category) verbal fluency ("fruits", 60 s); measures semantic fluency and lexical-semantic retrieval. | Verbal fluency - fruit names | 1,241 | education |
| GCSE | General Certificate of Secondary Education science exam (England, ~16 y); measures educational attainment in science (not a clinical test). | Science exam score | 1,703 | sex |
| Personality | Openness subscale of the International Personality Item Pool (IPIP; 5 self-report items); measures openness to experience. | Openness scale score | 2,137 | education |
| STAS | Trait-Anger scale of the State-Trait Anger Scale (STAS; self-report); measures trait anger. | Trait Anger score | 316 | sex |
| TMAS | Taylor Manifest Anxiety Scale (self-report questionnaire); measures manifest (trait) anxiety (reverse-scored: lower = more anxious). | Manifest Anxiety score | 523 | sex |

**Table 2.** Model complexity across OLS, GAM, and GAMLSS specifications. k is the number of predictors retained in the OLS model. EDF indicates how flexible the GAM age curve is; values close to 1 mean an almost linear effect, whereas higher values mean more curvature. sigma-ratio compares the largest and smallest residual standard deviation estimated by GAMLSS; values close to 1 mean that residual variability is approximately constant. The correlations are obtained by Pearson correlations between the Z-scores produced by each pair of models.

|  |  | Correlation between models |  |  |  |  |
| --- | --- | --- | --- | --- | --- | --- |
| Dataset | k | OLS vs.<br>GAM corr | OLS vs.<br>GAMLSS<br>corr | GAM vs.<br>GAMLSS | EDF | Sigma-ratio |
| Substitution (LDST) | 3 | 0.9973 | 0.9936 | 0.9962 | 9.9 | 1.32 |
| VLT | 5 | 0.9962 | 0.9945 | 0.9982 | 11.9 | 1.26 |
| Fluency | 2 | 1.0000 | 0.9717 | 0.9717 | 3.0 | 1.79 |
| GCSE | 1 | 1.0000 | 0.9999 | 1.0000 | 1.9 | 1.02 |
| Personality | 4 | 0.9997 | 0.9957 | 0.9960 | 4.3 | 1.32 |
| STAS | 0 | 1.0000 | 1.0000 | 1.0000 | 1.1 | 1.00 |
| TMAS | 1 | 0.9999 | 0.9796 | 0.9797 | 1.9 | 1.50 |

### 3.2 Comparison 1: each model reproduces the classic norms

All three regression models reproduced the classic Z-scores closely, with correlations between r = 0.964 and r = 1.000 (Figure 1; Table 3). The two accuracy metrics provide equivalent information once the scale issue was handled: residual RMSE was small relative to each score range, and the percentage of participants matching the classic Z within 0.25 was high across models. In the two heteroscedastic datasets, GAMLSS reached 100% agreement within 0.25 (Fluency, TMAS), consistent with its ability to model subgroup-specific variance. For practical interpretation, this first comparison shows that OLS, GAM, and GAMLSS can all reproduce established norms when the reference data are sufficiently informative.

**Table 3.**
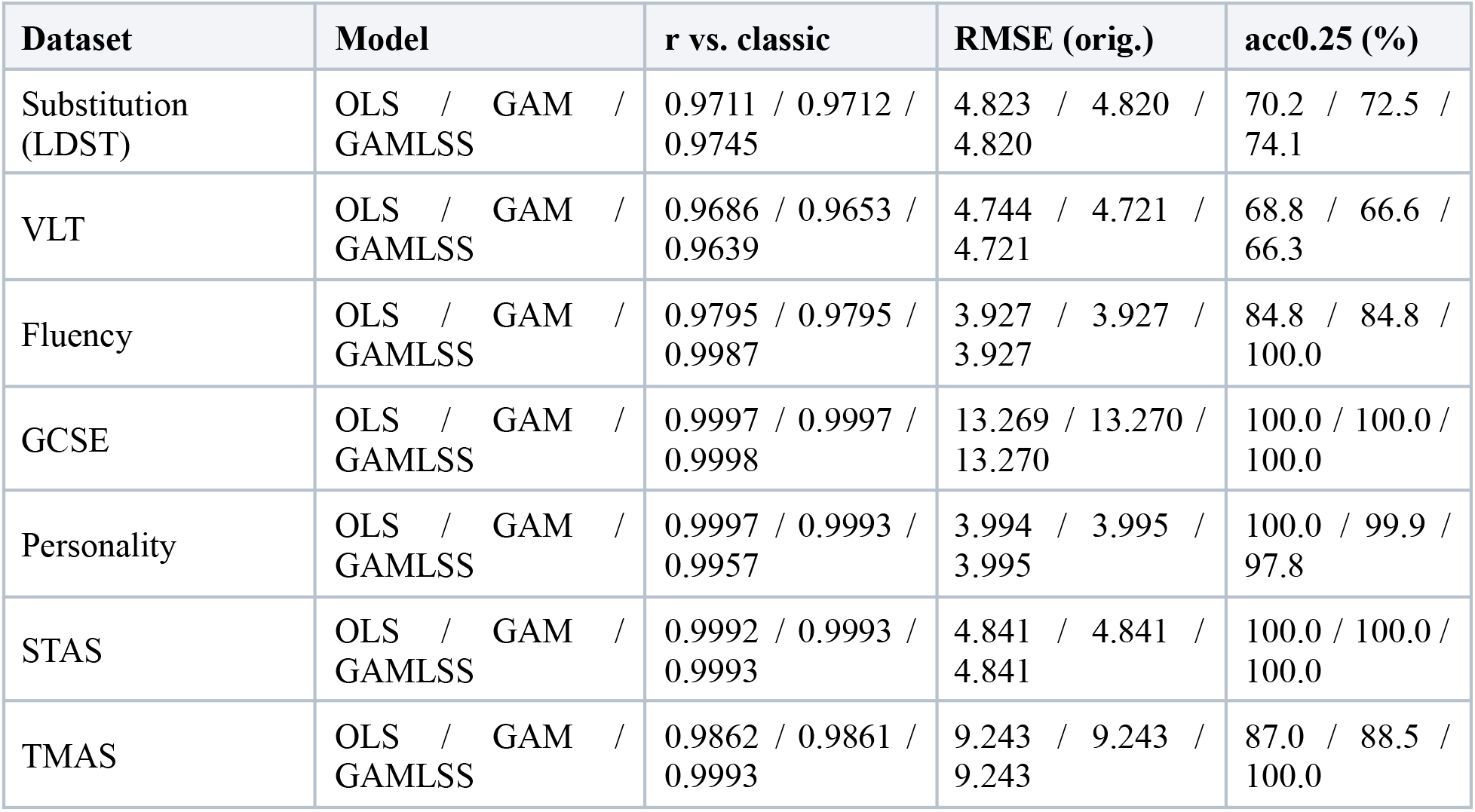
Agreement and prediction accuracy of OLS, GAM, and GAMLSS normative models. The three models provide high accuracy. GAMLSS gains under heteroscedasticity. The table shows correlation, prediction error, and same-classification agreement side by side. For each model and dataset, we provide the Pearson r with the classic Z, the residual RMSE on the original score scale, and the acc0.25 (ie., the percentage of participants whose standardized model-based Z agrees with the standardized classic Z within 0.25).

**Figure 1.**
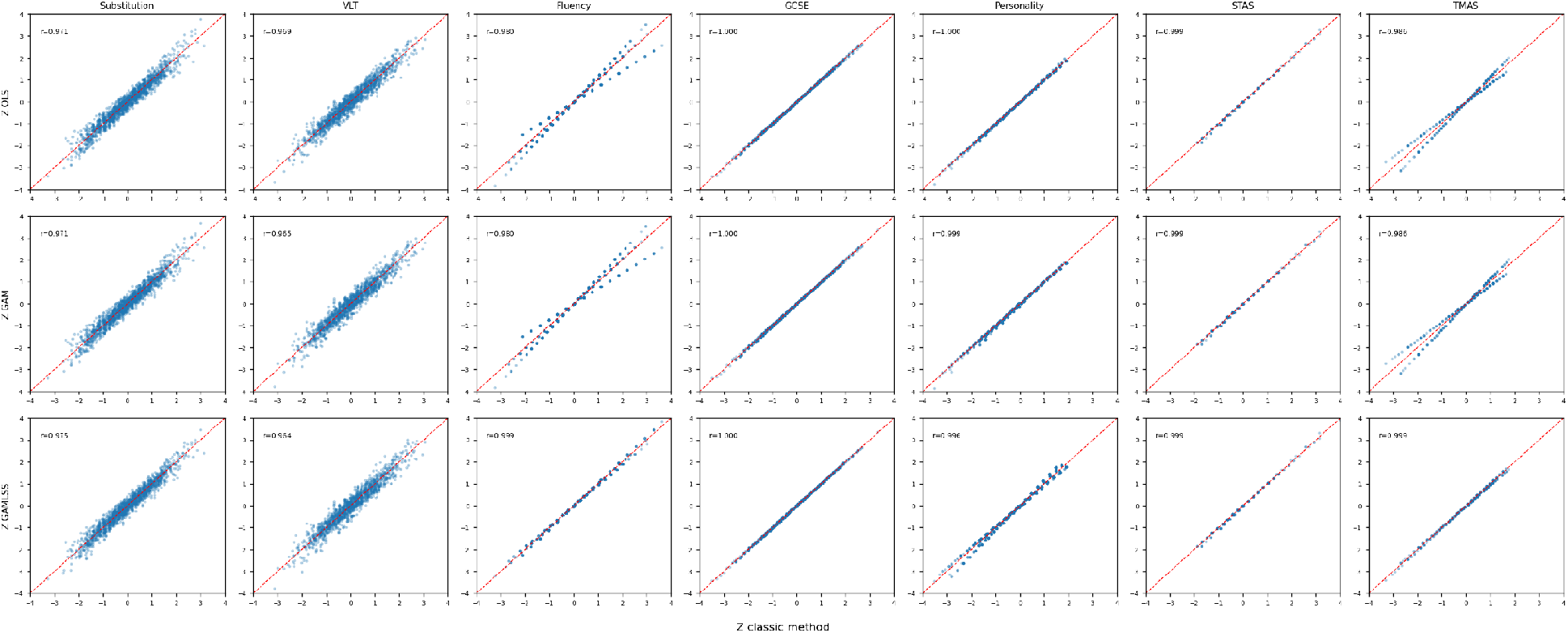
All regression models reproduce the classic norms. If the dots follow the diagonal, the model gives nearly the same individual interpretation as the traditional norm. Concordance of OLS, GAM, and GAMLSS Z-scores with the classic Z-score across the seven datasets. The GAMLSS row uses the bias-corrected, standardized Z-score; the dashed line is the identity line.

**Figure 2.**
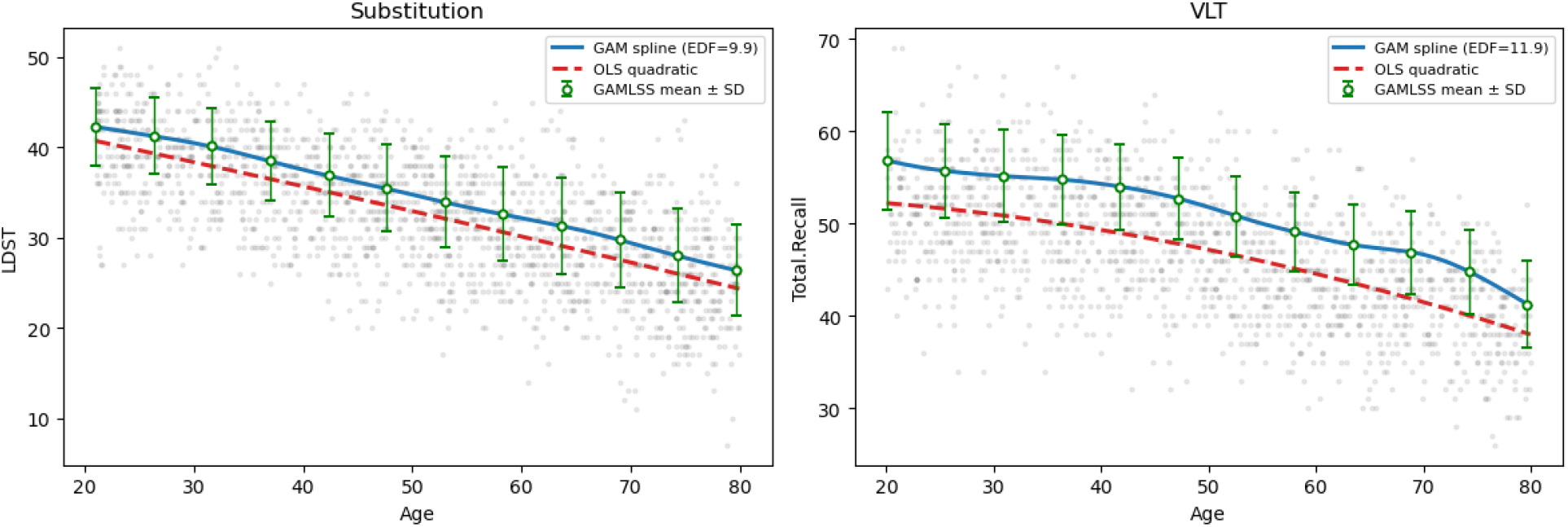
Examples of different model behaviors. For the two datasets with a continuous age covariate, three estimates of the age trend are represented: the GAM age spline (solid line), the OLS quadratic fit (dashed line), and the GAMLSS fitted mean with ±1 standard-deviation error bars (markers). The GAM spline and the GAMLSS mean follow the same age trend, and although the OLS quadratic is slightly stiffer, it does not materially change the central-tendency estimate. Crucially, the error bars display the conditional standard deviation estimated by the GAMLSS scale submodel — the component that OLS and GAM do not account for as both OLS and GAM assume constant variance. Modelling this scale explicitly is what allows GAMLSS to accommodate age-dependent dispersion in the response. Age spline EDF = 9.9 (LDST) and 11.9 (VLT).

### 3.3 Comparison 2: what GAM and GAMLSS add over OLS

OLS and GAM produced essentially identical Z-scores in every dataset (r = 0.996-1.000), even when the GAM allocated many effective degrees of freedom to the age spline. This indicates that the observed age-related nonlinearities were smooth enough to be captured by the quadratic term already included in OLS. A higher EDF means that the curve bends, but, importantly, it does not automatically mean that the simpler model fails.

GAMLSS diverged from OLS specifically in datasets with non-constant residual variance. The largest differences occurred for Fluency (OLS vs. GAMLSS r = 0.972, sigma-ratio = 1.79) and TMAS (r = 0.980, sigma-ratio = 1.50). In those datasets, GAMLSS agreed more closely with the classic norms than OLS did, because the classic method already computes a separate standard deviation within each subgroup. GAMLSS recovers that behavior explicitly by modeling sigma as a function of covariates, whereas OLS imposes a single constant sigma.

### 3.4 GAMLSS scale bias and its correction

The two-stage GAMLSS estimator carried the expected scale bias: the fitted log-variance was biased downward, which made the empirical standard deviation of raw GAMLSS Z-scores too large (1.79-2.24; Table 4). Adding the Harvey correction rescaled sigma as intended, but because it is a single global factor it did not change the OLS-GAMLSS Pearson correlations. We retain the correction for methodological transparency and standardize GAMLSS Z-scores before comparing them with classic or OLS Z-scores.

**Table 4.** Effect of the Harvey correction on GAMLSS Z-score scaling and model agreement. Overall, the correction changes the absolute spread of GAMLSS Z-scores but leaves correlations unchanged. Empirical standard deviation of GAMLSS Z before and after correction, and OLS-GAMLSS correlation before and after correction. Raw sd(Z) reflects absolute miscalibration of the two-stage GAMLSS sigma estimate; the Harvey constant (1.2704) rescales sigma but leaves standardized Z-score correlations unchanged.

| Dataset | sd(Z) raw | sd(Z) corr. | OLS-GAMLSS raw | OLS-GAMLS S corr. | sigma-ratio |
| --- | --- | --- | --- | --- | --- |
| Substitution (LDST) | 1.851 | 0.981 | 0.9936 | 0.9936 | 1.32 |
| VLT | 1.895 | 1.004 | 0.9945 | 0.9945 | 1.26 |
| Fluency | 2.131 | 1.129 | 0.9717 | 0.9717 | 1.79 |
| GCSE | 1.893 | 1.003 | 0.9999 | 0.9999 | 1.02 |
| Personality | 1.799 | 0.953 | 0.9957 | 0.9957 | 1.32 |
| STAS | 2.241 | 1.188 | 1.0000 | 1.0000 | 1.00 |
| TMAS | 1.785 | 0.946 | 0.9796 | 0.9796 | 1.50 |

## 4. Discussion

Across seven heterogeneous instruments created for neuropsychological and psychological assessment, OLS reproduced the classic Z-scores with correlations between 0.969 and 1.000 and produced Z-scores almost identical to GAM. Combined with transparency, interpretability, and modest data requirements, this makes OLS a suitable model for normative Z-score computation. Each coefficient has a direct meaning, which supports clinical trust in the normalization procedure (Mitrushina et al., 2005; Strauss et al., 2006; Sherman et al., 2022).

The very high OLS-GAM agreement shows that a flexible spline did not materially change the normative scores in these datasets. GAM would be expected to add value mainly when age effects are sharply nonlinear, for example under accelerated late-life decline that a low-order polynomial cannot follow (Der & Deary, 2006; Salthouse, 1996). In the present data that condition did not arise, so GAM is best regarded as a principled extension rather than a routine replacement for OLS.

GAMLSS differed from OLS precisely where residual spread was not constant, and there it improved fidelity to the classic norms. Conversely, when heteroscedasticity was weak, the scale model could slightly reduce agreement with the classic norm. From here, we can infer a practical rule: To use GAMLSS when there is evidence that variability changes across covariate profiles and enough data to estimate that change; otherwise, the constant-sigma assumption of OLS is preferable.

Here, we adopted the rank-based Z-score as the final score for all datasets, which sidesteps the normality assumption of the parametric Z-score without sacrificing concordance with the classic norms. This matters because bounded-scale instruments can produce skewed residuals owing to floor and ceiling effects. Sample size is not the central uncertainty here: regression-based norming can be efficient when designed well (Innocenti et al., 2023), and all seven public datasets were sufficiently large for the models fitted.

In the context of digital assessment, our work has shown that there is more than one valid strategy to derive normative data. Candidate models should first be benchmarked on public datasets and, when the reference sample is large enough for cell-based estimates to be meaningful, against the classic subgroup method. The point is not to rank methods as intrinsically better or worse, but to understand which model is appropriate for the structure that is actually observable in the data. OLS remains the baseline for transparent adjustment, GAM is useful when the conditional mean has a nonlinear structure, and GAMLSS is useful when residual variance changes across covariates. Thus, model choice should depend on the modeling question, sample size, covariates, and the amount of reliable information the dataset contains; complexity is justified only when it changes what we can responsibly say about an individual score.

Two key limitations should be noted. First, for our analyses here, only two of the seven datasets contain a continuous age covariate, which is where GAM and GAMLSS have the greatest potential to differ from OLS. Second, the GAMLSS implementation used a two-stage approximation rather than dedicated GAMLSS software. Future work should test the same framework in larger digital cohorts, datasets with stronger nonlinear age trajectories, and data-generating processes that include bounded or discrete outcomes, where raw-score regression and distributional modeling choices are especially important (Oltra-Cucarella et al., 2025).

To conclude, we fitted OLS, GAM, and GAMLSS to all seven reference datasets of the NormData package and assessed both their equivalence to classic subgroup norms and their agreement with one another. All three models reproduced the classic norms closely. OLS and GAM produced almost identical Z-scores, indicating that smooth age effects were already captured by a quadratic term, whereas GAMLSS diverged from OLS only under heteroscedasticity and improved fidelity to classic norms in those cases. These results support a pragmatic normative-model selection rule: begin with interpretable OLS, add GAM only for substantial nonlinear mean structure, and add GAMLSS only when residual variance depends on covariates.

## Data Availability

All datasets analyzed in this study are publicly available through the NormData R package (version 1.1; Van der Elst, 2024). The seven datasets analyzed were publicly available before the initiation of this study. No new data were collected for this study.

https://cran.r-project.org/web/packages/NormData/index.html

https://doi.org/10.32614/CRAN.package.NormData

## 5. Additional Information

### 5.1 Author Contributions

M.A.G.: Conceptualization; formal analysis; investigation; methodology; visualization; writing – original draft; writing – review and editing. P.R.R.: Data curation; validation. E.M.: Data curation; validation. C.S.B.: Validation; writing – review and editing. J.M.Corada: Data curation; validation. C.O.S.: Data curation; validation. I.F.d.P: Project administration; resources. M.R.L: Conceptualization; methodology; resources; supervision; writing – review and editing. J.M.Cortes: Conceptualization; formal analysis; methodology; supervision; writing – review and editing.

### 5.4 Use of AI-assisted tools

Claude (Anthropic) and Codex (OpenAI) were used for language editing. The authors reviewed all outputs and assumed full responsibility for the content of the manuscript.

